# Blood epigenome-wide association studies of suicide attempt in adults with bipolar disorder

**DOI:** 10.1101/2023.07.20.23292968

**Authors:** Salahudeen Mirza, Camila N. de Carvalho Lima, Alexandra Del Favero-Campbell, Alexandre Rubinstein, Natasha Topolski, Brenda Cabrera-Mendoza, Emese H.C. Kovács, Hilary P. Blumberg, Jenny Gringer Richards, Aislinn J. Williams, John A. Wemmie, Vincent A. Magnotta, Jess G. Fiedorowicz, Marie E. Gaine, Consuelo Walss-Bass, Joao Quevedo, Jair C. Soares, Gabriel R. Fries

## Abstract

Suicide attempt (SA) risk is elevated in individuals with bipolar disorder (BD), and DNA methylation patterns may serve as possible biomarkers of SA. We conducted epigenome-wide association studies (EWAS) of blood DNA methylation associated with BD and SA. DNA methylation was measured at > 700,000 positions in a discovery cohort of n = 84 adults with BD with a history of SA (BD/SA), n = 79 adults with BD without history of SA (BD/non-SA), and n = 76 non-psychiatric controls (CON). EWAS revealed six differentially methylated positions (DMPs) and seven differentially methylated regions (DMRs) between BD/SA and BD/non-SA, with multiple immune-related genes implicated. There were no epigenome-wide significant differences when BD/SA and BD/non-SA were each compared to CON, and patterns suggested that epigenetics differentiating BD/SA from BD/non-SA do not differentiate BD/non-SA from CON. Weighted gene co-methylation network analysis and trait enrichment analysis of the BD/SA vs. BD/non-SA contrast further corroborated immune system involvement, while gene ontology analysis implicated calcium signalling. In an independent replication cohort of n = 48 BD/SA and n = 47 BD/non-SA, fold-changes at the discovery cohort’s significant sites showed moderate correlation across cohorts and agreement on direction. In both cohorts, classification accuracy for SA history among individuals with BD was highest when methylation at the significant CpG sites as well as information from clinical interviews were combined, with an AUC of 88.8% (CI = 83.8-93.8%) and 82.1% (CI = 73.6-90.5%) for the combined epigenetic-clinical predictor in the discovery and replication cohorts, respectively. Our results provide novel insight to the role of immune system functioning in SA and BD and also suggest that integrating information from multiple levels of analysis holds promise to improve risk assessment for SA in adults with BD.

## Introduction

Suicide is a leading cause of death, with over 47,000 suicide deaths (SD) in the United States in 2021. Many more people attempt suicide each year, with an estimated 1,400,000 suicide attempts (SA) in the US in 2021 [1]. Improving detection of SA risk could be critical to reducing injuries, as well as intervening early to prevent SD. There have been various efforts to identify robust and easily obtainable clinical predictors. For example, in a study from our group with mood disorders subjects, a combination of risk factors including previous hospitalisations for depression, history of psychosis, cocaine dependence, and post-traumatic stress disorder (PTSD) has shown potential for classifying patients based on SA history [2]. However, such classification tools are still nascent, and a recent meta-analysis of canonical risk factors for SA and other suicidal thoughts and behaviours found that there are no strong longitudinal predictors [3].

Applying a multiple-levels-of-analysis approach may be key to understanding the aetiology of SA [4, 5] and identifying signs of risk in vulnerable populations. Integration of information across the biological and clinical domains, when taken together, could illuminate novel pathways and opportunities for intervention. Suicidal behaviour has been shown to have a biological component [6, 7]. Emerging genome-wide association studies (GWAS) are beginning to identify associated genes, but the variance explained is low [8, 9]. Beyond the contributions of the genotype itself, mechanisms which regulate gene expression may also be relevant. DNA methylation, the reversible addition of methyl groups to the DNA sequence, can modulate gene expression and ultimately shape larger biological networks [10]. Accordingly, DNA methylation alterations have been proposed as possible mediators between early life adversity and the development of psychopathology, including suicide risk [11]. Of note, some of the earliest work in psychiatric epigenetics demonstrated DNA methylation alterations related to childhood abuse and SD [12].

Patterns in DNA methylation may also capture genetic and environmental contributions to SA. Epigenome-wide association studies (EWAS), which attempt to surveille DNA methylation across the entire (epi)genome, improve upon many of the limitations of previous studies focused on specific candidate genes [9]. The majority of suicide-focused EWAS have considered SD, as opposed to a few which studied SA [13–15]. There are several indicators that the epigenetic basis may differ between SA and SD [9]. Aside from a variety of epidemiological differences [16], the existing EWAS of SD are predominantly based in brain tissue [9], which is generally inaccessible in living individuals. DNA methylation patterns vary by cell type and can differ between peripheral tissues and the brain [17]. In particular, understanding the contributions of peripherally-measured DNA methylation to SA *in vivo* would best inform preventive and interventional strategies, such as screening tools. Recently, a large blood EWAS of SA in U.S. military veterans identified three significant differentially methylated positions (DMPs) associated with SA, implicating nervous system-related alterations and overlapping DNA methylation patterns with a variety of risk traits [15]. Carrying this momentum forward, it is important to understand the associated peripheral DNA methylation patterns in other focused subgroups, including specific psychiatric diagnoses, given evidence that the molecular underpinnings of SA may differ across diagnostic populations [9].

Risk for SA is substantially elevated in individuals with bipolar disorder (BD), with 20-60% of individuals having at least one lifetime SA [18]. This elevated risk could be accounted for by patterns of shared biological vulnerability, with recent GWAS suggesting genetic correlation [19]. BD is also associated with other psychiatric comorbidities and early adversities which are known to drive suicide risk [20]. One of the challenges of the extant literature on epigenetics of suicidal behaviour is that a suicidal case group is often compared only to a non-psychiatric control group. A central limitation of this approach is that any pathophysiological alterations described in the cases cannot be ascribed to suicide *vs*. the elevated burden of psychopathology [9]. Therefore, there is a need to carefully study the epigenetics of SA using well-matched reference groups, such as comparing individuals with BD and a history of SA to individuals with BD without such history. Extending the study to also include non-psychiatric controls can inform on the extent to which SA-associated alterations are unique from BD-associated pathology (e.g., whether individuals with BD and SA are showing an exacerbation of BD-associated pathology). Results from EWAS of SA in BD could provide insights to the underlying biology, as well as provide possible diagnostic aids. Critically, this information from the biological domain, instead of being considered independently from the clinical domain, could be integrated with existing measures, such as clinical interviews [21].

In this study, we leveraged a richly phenotyped discovery cohort of individuals with BD stratified by lifetime history of SA, as well as non-psychiatric controls, to understand the underlying epigenetic vulnerabilities linked to SA. We conducted EWAS to identify differentially methylated positions (DMPs) and regions (DMRs) associated with BD and SA. We measured the association of these DNA methylation patterns with other traits and biological pathways. To address concerns of replicability, we attempted to validate our findings in an independent replication cohort of individuals with BD and SA. Finally, to demonstrate the potential of integrating information across multiple levels of analysis to inform risk assessment, we tested the ability of various models to predict SA history based on DNA methylation and clinical measures both independently and jointly in the discovery and replication cohorts.

## Method

### Discovery cohort

This sample has been previously described [22]. One-hundred and sixty-one adults with BD (79 BD/non-SA, 84 BD/SA) and 76 non-psychiatric controls without a lifetime history of SA (CON) were recruited at the Center of Excellence in Mood Disorders, Houston, TX. BD diagnosis was ascertained in the Structured Clinical Interview for DSM-IV Axis I Disorders (SCID-I) [23]. Lifetime history of SA was assessed with Columbia Suicide History Form (CSHF) [24]. Other demographic and clinical characteristics (e.g., substance use, previous hospitalisations, psychiatric comorbidities) were obtained by demographic questionnaire and clinical interview. Interviews were administered by trained evaluators and reviewed by a board-certified psychiatrist. Young Mania Rating Scale (YMRS) [25] and Montgomery-Asberg Depression Rating Scale (MADRS) [26] were administered for assessing manic and depressive symptomatology. Exclusion criteria for all participants included neurological disorders and traumatic brain injury, schizophrenia, developmental disorders, eating disorders, intellectual disability, and recent illicit drug use by urine drug screen. Exclusion criteria for CON included a history of any Axis I disorder in first-degree relatives or if they had taken a prescribed psychotropic medication at any point in their lives. The study protocol was approved by the local institutional review board (IRB), and informed consent was obtained from all participants at enrolment and prior to any procedure.

### Replication cohort

This sample has been previously described [22]. Ninety-five adults with BD (48 BD/SA, 47 BD/non-SA) were recruited through the IRB-approved Iowa Neuroscience Institute Bipolar Disorder Research Program of Excellence (BD-RPOE). There were no CON members in the replication cohort. Participants between 18 and 70 years with a confirmed SCID-I diagnosis of BD-I provided informed consent. History of SA and number of lifetime attempts were recorded with the Columbia Suicide Severity Rating Scale (C-SSRS) [24]. Demographic measures and clinical interviews, including YMRS and MADRS, were administered. Exclusion criteria included a history of loss of consciousness for more than 10 min, seizure disorder, brain damage or other neurological problems, coronary or cerebral artery disease, alcohol or drug dependence within the past 3 months, current pregnancy, or contraindication for magnetic resonance imaging.

### Methylation assay

All participants provided peripheral blood by venipuncture, which was stored in EDTA-containing vacutainers at -80 degrees Celsius. In the discovery cohort, DNA was isolated from the buffy coat using the DNeasy Blood & Tissue Mini Kit (Qiagen, Hilden, Germany). In the replication cohort, 1 mL of whole blood per sample was used with the Puregene Blood Kit with RNase A solution (Qiagen, Hilden, Germany). Elution Buffer CDB-02 (Kurabo Industries Ltd, Osaka, Japan) was used instead of DNA Hydration Solution. In both cohorts, five hundred nanograms of DNA were bisulfite-converted using the EZ DNA Methylation Kit (Zymo Research, Irvine, CA, USA). Genome-wide DNA methylation was measured using the Infinium EPICMethylation BeadChip version 1.0 (Illumina, San Diego, CA, USA), according to manufacturer’s instructions.

### Information for ancestry controls

In the discovery cohort, DNA was hybridised to the Infinium Global Screening Array v1.0 and v3.0 (Illumina, San Diego, CA, USA) to measure common genetic variation. Prior to principal components analysis, pruning was performed in PLINK v1.9 [27] with a window size of 200 variants, step size of 50 variants, and LD r^2^ threshold of 0.25. The first three principal components of genotyping data were extracted for each participant and retained as covariates for genomic ancestry. In the replication cohort, EPISTRUCTURE, which uses information from patterns in DNA methylation rather than genotyping [28], was used to compute the first three principal components used for ancestral adjustment. EPISTRUCTURE has been validated as an alternative to using genotyping information to capture ancestry [28].

### Data pre-processing, quality controls, and filtering

All pre-processing and analysis steps were performed in R version 4.2.0 with the package *minfi* [29]. Cross-reactive probes [30], probes with mean detection *p*-value > 0.01, probes with fewer than 3 beads in > 1% of samples, polymorphic probes [30], and probes located on the sex chromosomes were removed. Quality controls were conducted separately for each EWAS carried out. There were four EWAS performed: discovery cohort BD/SA vs. BD/non-SA (719,231 CpG sites), discovery cohort BD/SA vs. CON (725,943 CpG sites), discovery cohort BD/non-SA vs. CON (725,843 CpG sites), and replication cohort BD/SA vs. BD/non-SA (654,448 CpG sites). In the discovery cohort BD/SA vs. BD/non-SA EWAS, one member of the BD/SA group failed quality controls, reducing the size of the BD/SA group in that analysis to n=83.

### Statistical analysis

Methylation values were log-transformed (base 2) from beta-values to M-values [31]. Control for batch effects and technical variability was achieved by the functional normalisation method, which applied noob background correction, dye normalisation, and correction for the first 2 principal components of the internal control probes on the EPIC array to the M-values [32]. To address cell type heterogeneity, white blood cell count proportions (CD8+ T cells, CD4+ T cells, natural killer cells, B cells, monocytes, and granulocytes) were estimated [33]. DNA methylation-based smoking scores were estimated based on a validated set of 174 sites in the R package *EpiSmokEr* [34, 35].

### EWAS for identification of DMPs

In both cohorts, epigenome-wide DMPs were measured with linear models adjusted for age, sex, white blood cell count proportions, DNA methylation smoking scores, and the first three ancestral principal components, in the R package *limma* [36]. Results for each EWAS were adjusted for false discovery rate (FDR) at significance threshold of *q* < 0.05 [37]. FDR-significant DMPs were annotated to the UCSC Genome Browser hg19 reference genome using the *CruzDB* Python implementation [38]. A sensitivity analysis of the BD/SA vs. BD/non-SA EWAS included further statistical adjustment for YMRS and MADRS total scores. The log2 fold changes were correlated across the discovery cohort EWAS (pairwise) for the FDR-significant DMPs, as well as the DMPs with nominal *p*< 0.001, from the BD/SA vs. BD/non-SA EWAS. This analysis primarily intended to better inform whether the underlying alterations differentiating BD/SA from BD/non-SA also differentiated BD/non-SA from CON.

### Blood-brain correlation of DMPs

The IMAGE-CpG tool, which provides within-person cross-tissue correlations for CpG sites measured on the EPIC BeadChip [17], was consulted to provide insight to the potential similarity of methylation levels at the discovery cohort FDR-significant DMPs between blood (measured in this study) and brain. The *rho* correlation coefficient for within-subject blood and brain methylation concordance was extracted for each FDR-significant DMP. The magnitudes of the coefficients were also averaged across the six DMPs to provide a broader sense of the generalizability of the blood-based results to brain methylation.

### Identification of DMRs

In all discovery and replication cohort EWAS, DMRs were identified using the *comb-p* algorithm in Python 2.7, a method based on autocorrelations between all probe *p*-values from the DMP analysis and subsequent combination as broader DMRs [39]. The following parameters were specified: seed-*p* value 1 x 10^-4^, minimum of 2 probes, and sliding window 500 base pairs. DMRs which passed a Šidák corrected *p*-value <0.05 were considered significant.

### Genomic location, trait, and pathway enrichment analyses of DMPs

The EWAS Toolkit [40] was used to test the DMPs from each EWAS with nominal *p* < 0.001 for enrichment for genomic locations (e.g., intergenic, gene body, etc), traits, and gene ontology (GO) pathways. The background used were probes on the EPIC BeadChip.

### Weighted gene co-methylation network analysis (WGCNA)

WGCNA was only performed for the BD/SA vs. BD/non-SA EWAS in the discovery cohort. Functionally normalised CpG probes were filtered based on a nominal *p*-value of < 0.05 and location at transcriptional start site (TSS) (n = 19,403 probes). The WGCNA R package was used to create co-methylated modules [41]. Soft power threshold of 9 was selected according to the criterion of approximate scale-free topology (R_signed_^2^ > 0.90). Eigengenes for each module were tested for association with SA in BD by logit generalised linear models including age, sex, white blood cell count proportions, smoking scores, and the first three ancestral genomic principal components as covariates, and *p*-values were adjusted for FDR by Benjamini-Hochberg procedure. GO analysis with the genes annotated to all CpG-sites comprising each module was performed using the EWAS Toolkit. The EWAS Toolkit has a maximum of 5,000 probes, so the first 5,000 were taken for the turquoise module. Probes related to hub genes were identified by module membership > 0.85 and gene significance > 0.30, with annotation of genes accomplished using *CruzDB* Python implementation.

### Statistical comparison across discovery and replication cohorts

At the FDR-significant DMPs in the discovery cohort, agreement in fold change direction was checked in the replication cohort. The log2 fold changes of the DMPs at *p* < 0.001 from the BD/SA *vs*. BD/non-SA contrast in the discovery cohort were correlated across the discovery and replication cohorts.

### Development of a clinical-epigenetic predictor of SA history

DMP methylation beta-values and six relevant clinical correlates (age, sex, number of previous hospitalisations for depression, history of psychosis, cocaine dependence, and PTSD) were tested to predict SA history among individuals with BD. The selected clinical correlates were the most important factors associated with SA in a previous machine learning analysis among individuals with mood disorders [2]. In individuals with BD from both cohorts, generalised linear models were fitted to predict SA history from (i) the beta-values from the six discovery cohort FDR significant DMPs alone, (ii) clinical correlates alone, and (iii) beta-values and clinical correlates together. Area under the curve (AUC) of the receiver operating curve (ROC) was used to evaluate the predictive performance of the models in R package pROC [42]. Additionally, pairwise differences in AUC among ROC curves were statistically tested using the DeLong test for two correlated ROC curves within cohorts.

## Results

### Demographic and clinical characteristics of the discovery and replication cohorts

Detailed information about the discovery and replication cohorts can be found in **Table 1**.

**Table 1.**
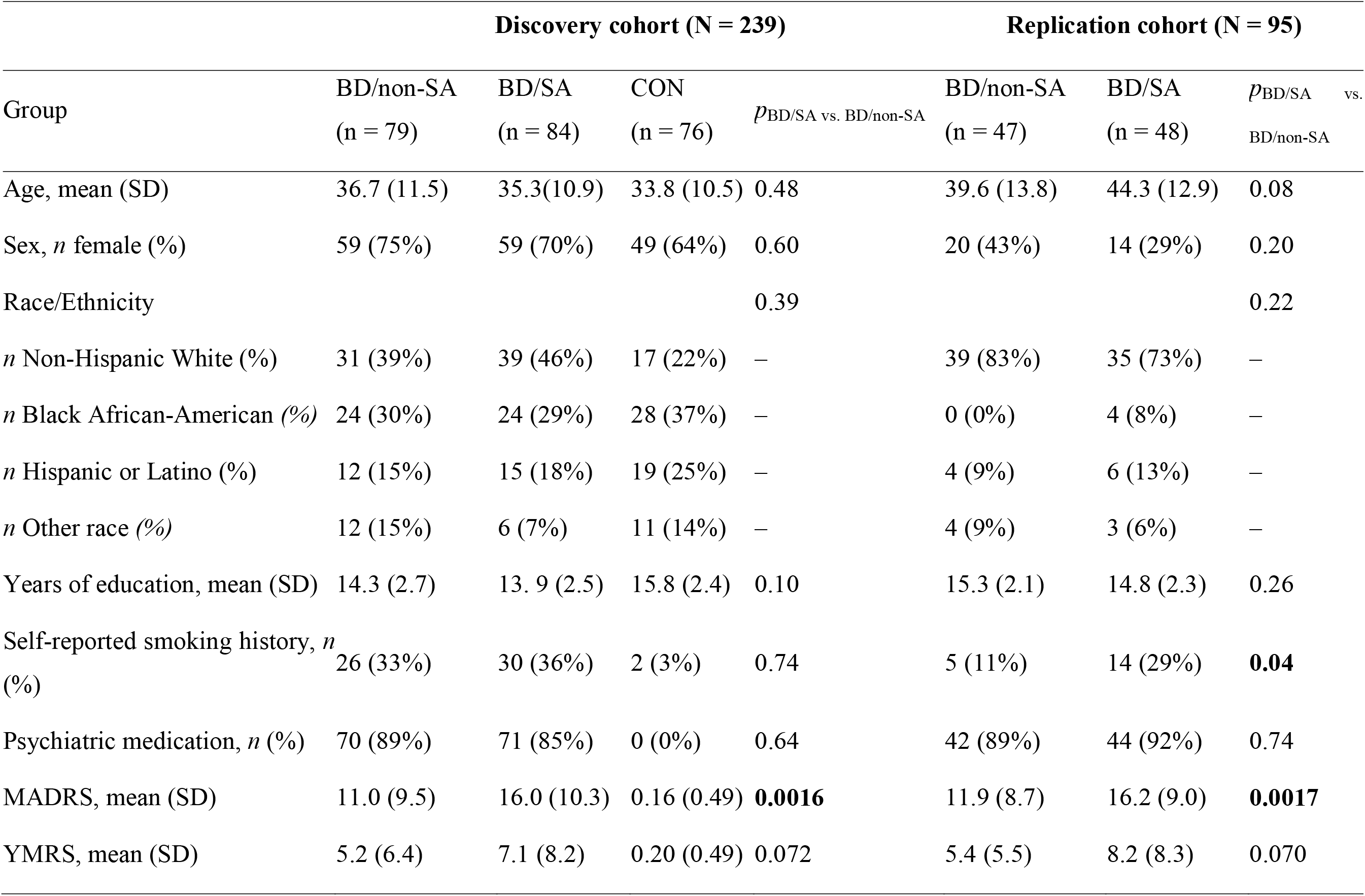
Demographics. Demographic and clinical characteristics of the discovery and replication cohorts Note on Table 1: p-values for quantitative variables were calculated using either t-test or Mann-Whitney U-test, depending on the result of a Shapiro-Wilk test for normality. p-values for categorical variables were calculated using the Fisher test. BD - bipolar disorder; CON - controls; MADRS - Montgomery-Asberg Depression Rating Scale; SA - suicide attempt; SD - standard deviation; YMRS - Young Mania Rating Scale.

### Discovery cohort EWAS identifies six DMPs between BD/SA and BD/non-SA

The discovery cohort EWAS identified six DMPs between BD/SA and BD/non-SA after FDR correction (*q* < 0.05) (**Figure 1 and Table 2**). The leading site was annotated to the *CXCL8* gene, which encodes the interleukin-8 cytokine. Two sites were hypomethylated in BD/SA: cg20244265 (*CXCL8*) and cg15653194 (*LFNG*). Four sites were hypermethylated in BD/SA: cg11476866 (*CD300LG*), cg20242392 (*DGKI*), cg25876840 (*CD300LG*), and cg00306112 (*LINC01494*) (**Table 2**). For all CpG site results at nominal (unadjusted) *p* < 0.001, see **Supplementary Table S1.** After sensitivity adjustment for YMRS and MADRS scores, fold changes at these six sites remained consistent and nominal *p*-values were all < 1 x 10^-5^, though none of the six sites survived the FDR correction **(Supplementary Table S2).**

**Figure 1.**
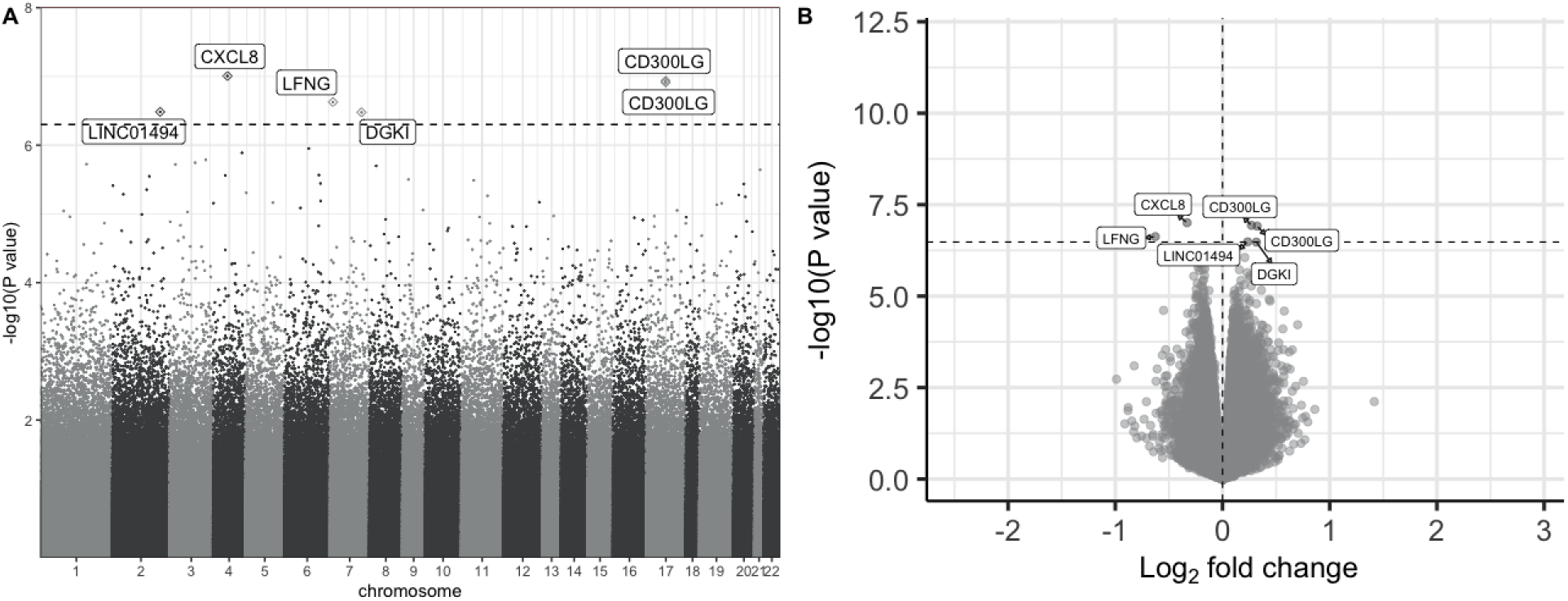
Results from the discovery cohort epigenome-wide association study of BD/SA (n=79) vs. BD/non-SA (n=83), demonstrated as **A)** Manhattan plot of significance by chromosomal location and **B)** Volcano plot of significance against log2 fold change, where positive and negative log2 fold changes indicate hypermethylation and hypomethylation in the BD/SA group, respectively. In both plots, the horizontal dotted line demarcates the FDR adjusted *q* < 0.05 threshold, and FDR-significant CpG sites are labelled based on their annotated genes. BD - bipolar disorder; FDR - false discovery rate; SA - suicide attempt.

**Table 2.** Differentially Methylated Positions. Six CpG sites differentially methylated between BD/non-SA and BD/SA at FDR *q* < 0.05 in the discovery cohort (n = 162).

| chr | Probe ID | Log2 FC | p | FDR q | Gene annotation | Gene distance (bp) | Gene feature |
| --- | --- | --- | --- | --- | --- | --- | --- |
| 4 | cg20244265 | -0.33 | $9.86 \times 10^{-8}$ | 0.03 | *CXCL8* | -22861 | Intergenic |
| 17 | cg25876840 | 0.27 | $1.16 \times 10^{-7}$ | 0.03 | *CD300LG* | -4066 | Intergenic |
| 17 | cg11476866 | 0.32 | $1.22 \times 10^{-7}$ | 0.03 | *CD300LG* | -4128 | Intergenic |
| 7 | cg15653194 | -0.63 | $2.36 \times 10^{-7}$ | 0.04 | *LFNG* | 0;0 | Exon+5'UTR; Intron |
| 2 | cg00306112 | 0.23 | $3.28 \times 10^{-7}$ | 0.04 | *LINC01494* | 0 | nc_intron |
| 7 | cg20242392 | 0.32 | $3.33 \times 10^{-7}$ | 0.04 | *DGKI* | 0;0 | Intron; Intron+5'UTR |

5'UTR - 5' untranslated region; BD - bipolar disorder; chr - chromosome; FC - fold change; FDR - false discovery rate; SA - suicide attempt.

The EWAS between BD/SA and CON found no DMPs at FDR correction *q* < 0.05 (full results at nominal *p* < 0.001 in **Supplementary Table S3**). In addition, the EWAS between BD/non-SA and CON also found no DMPs at FDR correction q < 0.05 (full results at nominal *p* < 0.001 in **Supplementary Table S4**, **Supplementary Figure S1**). Genomic inflation factor lambda was 1.20 for BD/SA vs. BD/non-SA, 0.90 for BD/SA vs. CON, and 1.21 for BD/non-SA vs. CON, suggesting no major issues of population stratification (**Supplementary Figure S2).**

At the six CpG sites which passed *q* < 0.05 in the BD/SA *vs*. BD/non-SA analysis, fold change direction was generally inconsistent among the three contrasts (BD/SA *vs.* CON, BD/SA *vs.* BD/non-SA, BD/non-SA *vs.* CON), suggesting noncontinuous changes between severity groups (**Supplementary Table S5, Supplementary Figure S3**). For the DMPs with *p* < 0.001 in the BD/SA vs. BD/non-SA analysis, the correlations among log2 fold changes between the BD/SA vs. BD/non-SA and BD/SA vs. CON contrasts were *r* = 0.76 (*p* < 2.2 x 10^-16^, df = 2,050), with proportion agreeing in fold change direction being 85% (95% CI = 84-87%). The correlation of the fold changes between the BD/SA vs. BD/non-SA and BD/non-SA vs. CON contrasts had an *r* = -0.84 (*p* < 2.2 x 10^-16^, df = 2,056), with proportion agreeing in fold change direction being 5% (95% CI = 4-6%). Finally, the correlation of fold changes between the BD/SA vs. CON and BD/non-SA vs. CON contrasts had an *r* = -0.36 (*p* < 2.2 x 10^-16^, df = 2,045), with proportion agreeing in fold change direction being 20% (95% CI = 18-22%) (**Supplementary Table S6, Supplementary Figure S4**). Twenty CpG probes passed *p* < 0.001 in both the BD/SA vs. BD/non-SA and BD/SA vs. CON analyses, as did 60 CpG probes in both the BD/SA vs. BD/non-SA and BD/non-SA vs. CON analyses, and 147 in the BD/SA vs. CON and BD/non-SA vs. CON analyses (listed in **Supplementary Table S7**).

### Blood-brain correlation of discovery cohort DMPs

Magnitudes of the blood-brain correlations for the six FDR-significant DMPs in the BD/SA vs. BD/non-SA contrast ranged from rho_abs_ = 0.01 (*CXCL8*) to rho_abs_ = 0.49 (*LFNG*), with a mean average of rho_abs_ = 0.29 and five of the six DMPs of magnitude greater than rho_abs_ = 0.20 (**Supplementary Table S8**).

### DMRs in the discovery cohort

In the discovery cohort, seven DMRs were identified between BD/SA and BD/non-SA after Šidák correction (Šidák *p* < 0.05) (**Supplementary Figure S5, Table 3**). The DMRs were annotated to the genes *TRIM40, RNF14, C14orf93, ETFBKMT, CD300LG, LOC728392,* and *HIVEP3.* There were no Šidák significant DMRs in the discovery cohort analyses of DMPs from BD/SA vs. CON and BD/non-SA vs. CON.

**Table 3.**
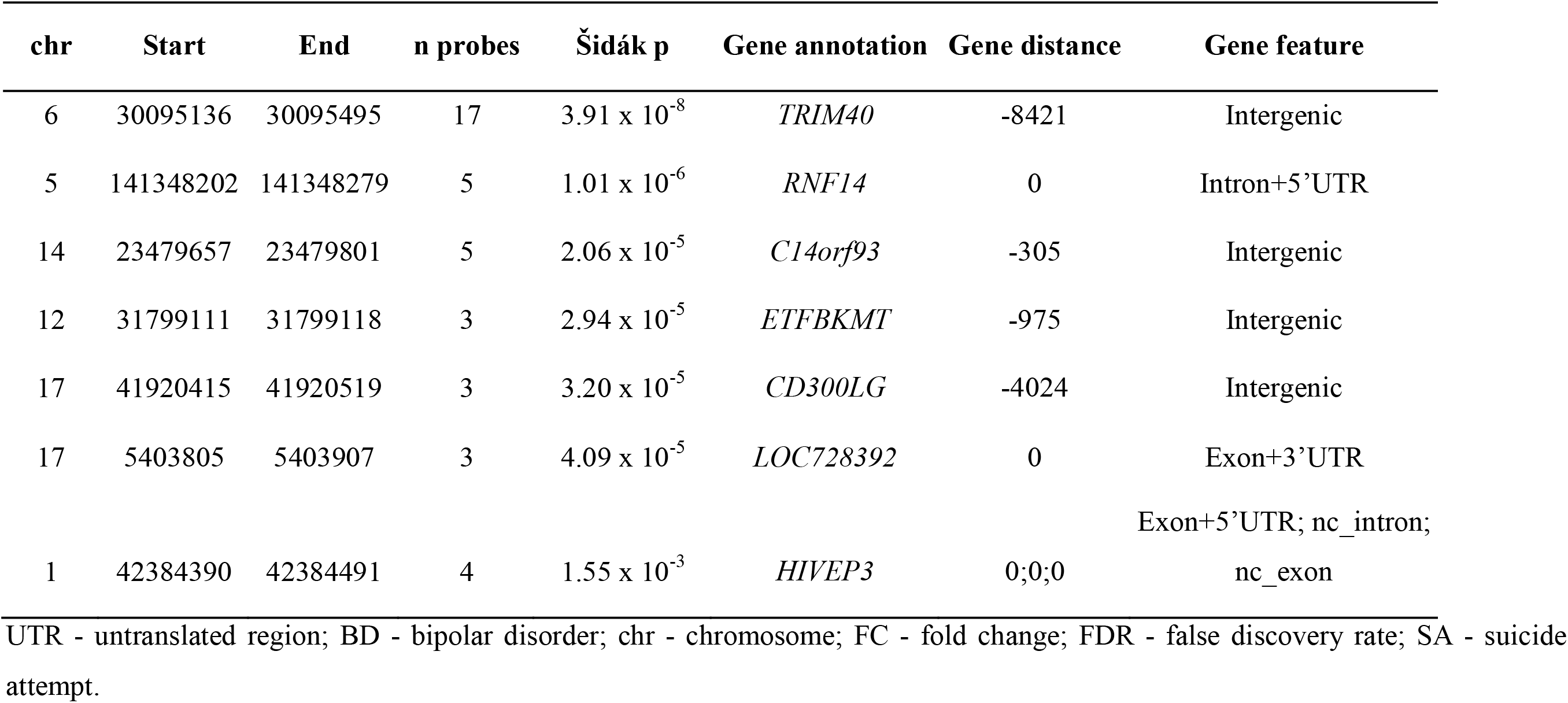
Differentially Methylated Regions. Seven regions differentially methylated between BD/non-SA and BD/SA at Šidák’s *p* < 0.05.

### Genomic location, trait and pathway enrichment analyses of DMPs in the discovery cohort

In the discovery cohort BD/SA vs. BD/non-SA EWAS, the DMPs were primarily concentrated in OpenSea (OR = 1.53) and intergenic (OR = 1.32) genomic locations. Top traits were especially enriched for inflammation-related traits. There was also trait enrichment for psychiatric and neurodevelopmental traits, as well as indicators of early life stress. Calcium signalling was implicated in the GO results. The results for the discovery cohort BD/SA vs. BD/non-SA EWAS can be viewed in **Figure 2** and **Supplementary Tables S9-S11.**

**Figure 2.**
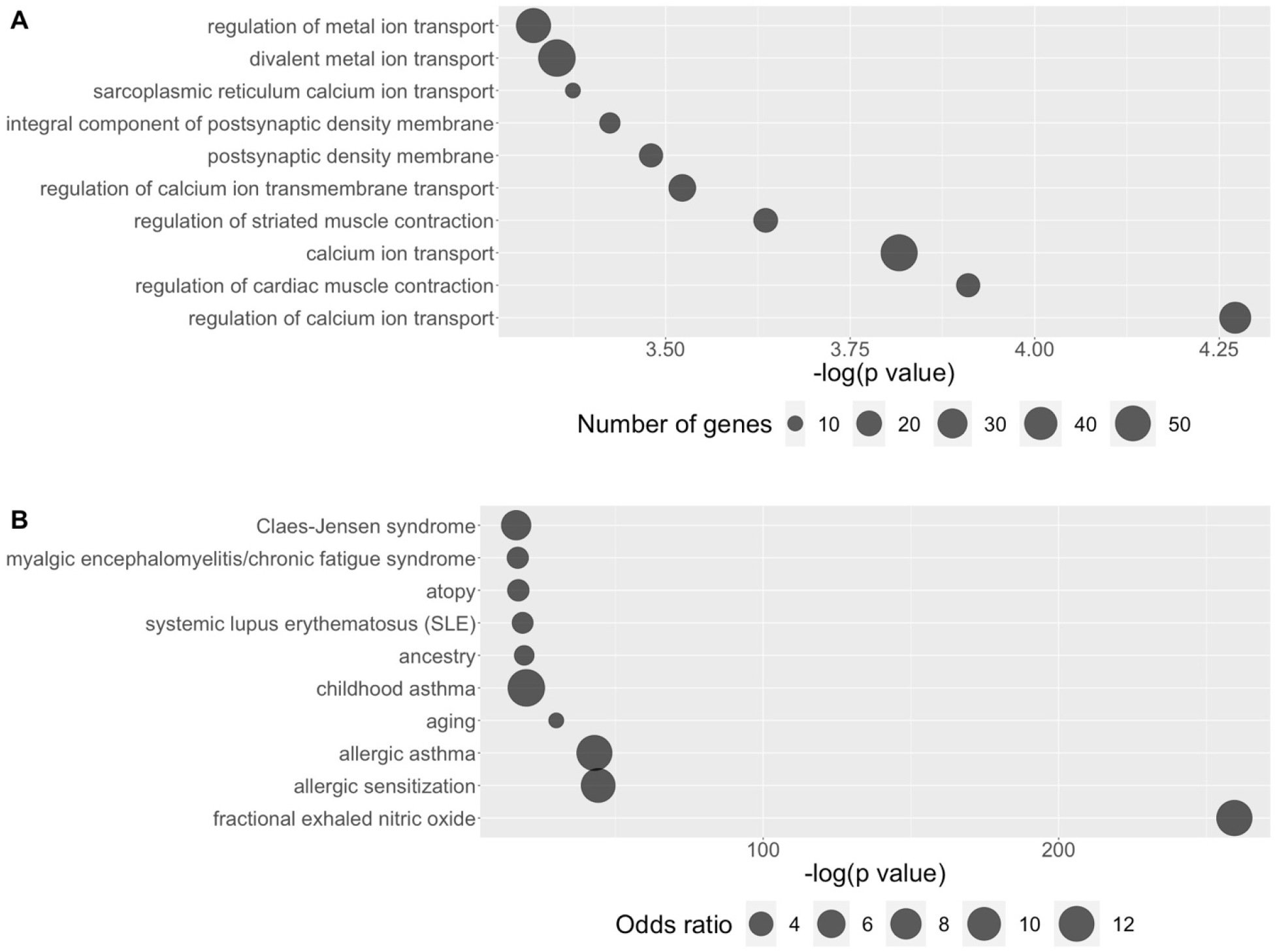
Gene ontology and trait enrichment results for the discovery cohort BD/SA vs. BD/non-SA EWAS using the DMPs at nominal p < 0.001. **A** shows the top ten gene ontology pathways ordered by -log(p value), with the number of genes assigned to each pathway represented by the size of the point. **B** shows the top ten traits ordered by -log(p value), with the odds ratio for each trait represented by the size of the point. BD - bipolar disorder; DMP – differentially methylated position; EWAS - epigenome-wide association study; SA - suicide attempt.

Enrichment for genomic locations, traits, and GO processes for the BD/SA vs. CON and BD/non-SA vs. CON EWAS can be seen in **Supplementary Figures S6-S7,** and **Supplementary Tables S9-S11**). There were no overlapping significant GO processes between the three contrasts.

### WGCNA in the discovery cohort

The WGCNA analysis returned six modules (blue, turquoise, brown, pink, black, green), with a mean of 2,947 probes per module and a range from 188 probes (pink) to 7,237 probes (turquoise) (**Supplementary Figure S8**). Unassigned CpG-sites (n=1,718) were clustered in a “grey” module, which was not considered for further analyses. All six modules were significantly associated with SA history in individuals with BD, led by the brown module (all *q* < 0.05) **(Supplementary Table S12**).

The assignment of probes to modules, including module membership and gene significance, can be found in **Supplementary Table S13.** The same information after a filter for hub genes is presented in **Supplementary Table S14.** Significant processes from the GO enrichment analyses performed with CpG probes in each module are shown in **Supplementary Table S15**. Of note, the top-ranked module, module brown, was substantially enriched for immune-related processes, including immune system process, immune response, and immune effector process (**Supplementary Table S15**). Relevant hub genes for this module included *RUSC1-AS1, BTG3-AS1, IQCE, SLC26A8, MIR6840, STAG3L5P-PVRIG2P-PILRB,* and *SSBP3* (for other module hub genes see **Supplementary Table S14**).

### Replication cohort validation of discovery cohort DMPs, DMRs, and enrichment analyses

In the replication cohort DMP analysis, no epigenome-wide significant CpG sites were identified at the FDR adjustment threshold of *q* < 0.05 (full results at nominal *p* < 0.001 in **Supplementary Table S16)**. Replication cohort log2 fold changes and *p*-values for the 6 discovery cohort epigenome-wide significant CpG sites are reported in **Supplementary Table S17,** with fold change direction being consistent at four of the six sites. None of the sites replicated at a nominal *p* < 0.05. Additionally, one of the sites (cg20242392, *DGKI*) failed quality control in the replication cohort so was not included in the DMP analysis. The replication cohort DMR analysis revealed no Šidák significant DMRs.

For the five discovery cohort FDR-significant CpG sites which survived filtering in the replication cohort, the correlation of log2 fold changes across cohorts was *r* = 0.60 (*p* = 0.28, df = 3). For the DMPs with *p* < 0.001 in the discovery cohort, the correlation of log2 fold changes across cohorts was *r* = 0.10 (*p* = 2.0 x 10^-5^, *df* = 1,956) (**Supplementary Table S18, Supplementary Figure S9)**. Additionally, the proportion of DMPs *p* < 0.001 which agreed in fold change direction across cohorts was 51% (95% CI = 49-53%).

Information on genomic locations, trait enrichment, and GO processes in the replication cohort BD/SA vs. BD/non-SA EWAS is available in **Supplementary Tables S19-S21)**. Some immune system-related traits and pathways arose in enrichment and GO analyses (**Supplementary Tables S20-S21)**. Several replication cohort significant traits overlapped with those from the discovery cohort BD/SA vs. BD/non-SA analysis, including inflammation-related diseases (e.g., perinatally-acquired HIV, Crohn’s disease) and indications of stress (e.g., preterm birth, maternal smoking). However, there was no overlap among GO terms across the discovery cohort and replication cohort contrasts.

### Performance of the clinical-epigenetic predictor of SA history among individuals with BD in the discovery and replication cohorts

Performance in both cohorts is presented in **Figure 3**. In the discovery cohort, the AUC was 83.7% (95% CI = 77.5-89.9%) for classification based on DMP methylation alone, 77.9% (95% CI = 70.6-85.3%) for classification based on clinical measures alone, and 88.8% (95% CI = 83.8-93.8%) for classification on combined methylation and clinical measures. In the replication cohort, the AUC was 67.6% (95% CI = 56.6-78.5%) for classification based on DMP methylation alone, 77.6% (95% CI = 68.2-86.9%) for classification based on clinical measures alone, and 82.1% (95% CI = 73.6-90.5%) for classification on combined methylation and clinical measures. DeLong’s test revealed that in both cohorts, classification based on DMP methylation significantly improved when clinical measures were included; though, adding DMP methylation to clinical measures only significantly improved classification in the discovery cohort (**Supplementary Table S22**).

**Figure 3.**
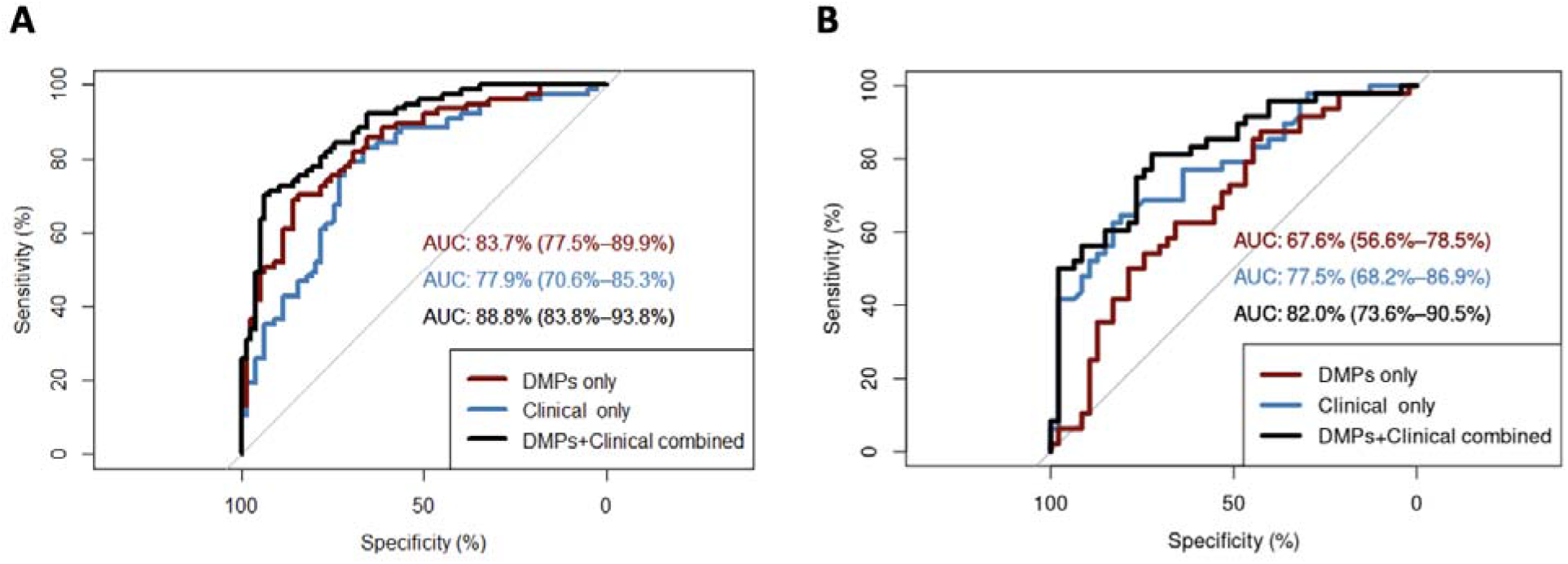
Receiver operating curves (ROC) for generalised linear models predicting SA history among individuals with BD, based on beta-values of the six FDR-significant discovery cohort DMPs (“DMPs only”), six clinical correlates of SA history in mood disorder (“Clinical only”), and the beta-values and clinical correlates combined (“DMPs+Clinical combined”). AUC of the ROC are presented with 95% confidence intervals. In **A**, ROC are presented for prediction of SA history in the discovery cohort (n=162). In **B,** ROC are presented for prediction of SA history in the replication cohort (n=95). AUC - area under the curve; DMP - differentially methylated positions; FDR - false discovery rate; SA - suicide attempt.

## Discussion

Considering the heightened risk for SA in BD, we conducted a series of EWAS of lifetime history of SA and BD. Further, we leveraged an independent replication cohort of individuals with BD to attempt to validate our findings and test a peripheral methylation biomarker of SA history within BD. In our discovery analysis, we identified six CpG sites and seven genomic regions which were significantly differentially methylated between individuals with BD with and without history of SA at stringent multiple testing thresholds. Importantly, the average blood-brain correlation for methylation at the CpG sites was low to moderate (average rho_abs_ = 0.29), suggesting that the identified patterns in DNA methylation may be specific to blood.

The top DMP was annotated to *CXCL8*, which encodes for the interleukin 8 cytokine and is a well recognized participant in the systemic immune response [43]. Further, gene *CD300LG*, which was mapped to two of the top DMPs and also captured by the DMR analysis, is also involved in immune system function, via modulation of cytokine-induced killer cells [44]. *LFNG*, annotated to one of the DMPs, is part of the Notch pathway, which has been implicated in neuro-inflammation and BD [45]. From the DMR analysis, *TRIM40* has recently been associated with inflammation in the context of inflammatory bowel disease [46], *RNF14* regulates mitochondrial and immune-related functioning [47], and *HIVEP3* regulates the development of innate-like T cells [48]. Further, a number of significantly associated traits from enrichment analysis were also associated with immune dysfunction. In addition, the top-ranked module from the WGCNA analysis was enriched for immune responses.

Taken together, these results suggest that alterations in immune-related genes in individuals with BD and SA history could provide an epigenetic mechanism for previously observed peripheral immune activation in SA [49–51]. The patterns of immune dysregulation in adults with BD and a history of SA may be attributable to increased exposure to environmental stressors [52]. Whether these alterations are simply correlates of a more stressful life or causal contributors to suicide risk remains to be seen, though it is possible that immune system changes lead to mood changes [53]. There is also the possibility that genetic influences are involved. Of note, the implications of the direction of methylation at these genes remain to be explored, ideally by concerted measurement of both DNA methylation and gene expression in follow-up work. At the functional pathway level, calcium signalling emerged as another possible pathway related to SA history among individuals with BD. In a previous study of BD and SD in post-mortem brain tissue, calcium signalling was similarly implicated [54]. The candidate gene *CACNA1C*, a calcium channel subunit, has previously been associated with BD and SA in both genetic and epigenetic analyses [55–58]. There were a number of significant GO processes related to neural functioning as well, though the direct effects are unclear as this study was conducted in peripheral blood.

Within the discovery cohort, we found evidence that the epigenetic pathophysiology differentiating BD/SA from BD/non-SA is not an exacerbation of pathology differentiating BD/non-SA from CON, as supported by the strong negative correlation of log2 fold changes and minimal percentage agreement in fold change direction at the nominally significant DMPs. This suggests that there is a specific epigenetic basis to SA in BD above and beyond the contributions of BD pathology in the absence of SA. Lack of FDR-significant results in the other contrasts could be partially due to the greater number of probes surviving quality controls, leading to a more stringent multiple comparisons correction.

In the independent replication cohort, we observed general consistency in the direction of fold change at the FDR-significant sites. Though none of the discovery cohort FDR-significant sites replicated at nominal *p* < 0.05 in the replication cohort, this could be due to a lack of statistical power in the replication cohort. Lack of statistical power could also be responsible for weaker correlation of fold changes across the discovery cohort nominally significant sites and limited convergence across GO processes, though there was some overlap in trait enrichment. An alternative effort to leverage the replication cohort was by inspecting the AUC of the ROC classification curve based on DMP methylation and clinical interviews. In both cohorts, methylation at the FDR-significant DMPs alone performed better than chance at predicting SA history among individuals with BD. Combining information on DNA methylation with information from clinical interviews performed best in both cohorts. Thus, patterns in DNA methylation could be used to augment existing risk prediction models and improve their performance. The multiple-levels-of-analysis approach to understanding SA risk in BD promises improvements to prevention and possible intervention [21].

There are several key limitations to consider in the interpretation of the study findings. As SA was ascertained retrospectively as a lifetime history, it is unclear how relevant this epigenetic signal may be for future SA risk. As some risk associated with suicidal behavior may be temporally enduring [12], there is the possibility that the EWAS are capturing some trait-like risk. However, there is also the possibility that, alongside the psychological consequences of SA [59], there may be concomitant biological sequelae. High rates of psychiatric medication in both cohorts render it difficult to isolate biological correlates of psychopathology from those of medication. In both cohorts, the EWAS were likely limited by statistical power constraints. Enrichment analyses and use of the replication cohort as a test sample for the AUC analyses were attempts to make the most of these limitations. As the EWAS were performed in peripheral blood, the immune-related findings may not extend to the brain (i.e., neuroinflammation), though this will have to be tested more directly in concerted follow-up studies. Finally, interpretation of DMPs, DMRs, and enrichment results relies on the assumption that DNA methylation alterations lead to changes in gene expression, although future studies incorporating gene expression measurements are needed to directly test this at relevant sites.

## Conclusion

In a series of EWAS, we investigated the epigenetic underpinnings of SA history in BD and provided replicable evidence that alterations in DNA methylation are especially concentrated at immune-relevant genes and related to systemic immune dysfunction. Though multilevel longitudinal research is needed to clarify the associated processes, we propose that these epigenetic modifications may be involved in the development of the atypical immune functioning which is implicated in suicide risk. Additionally, we demonstrate that epigenetic information could be useful for prediction of suicide risk and further that the greatest prediction accuracy arises when both epigenetic and clinical sources of information are considered together.

## Data Availability

All data produced in the present study are available upon reasonable request to the authors.

## Acknowledgments

This study was funded by the National Institute of Mental Health (NIMH: K01 MH121580 to GRF, R01MH125838 to VAM and JAW, and R01MH111578 to VAM and JAW), the American Foundation for Suicide Prevention (AFSP, YIG-0-066-20 to GRF), and the Carver Foundation. SM was funded by the GradSURP Summer Research Fellowship programme in the McGovern Medical School in 2021 and 2022. SM received funding from the Institute for Behavioral Genetics at the University of Colorado Boulder to attend the International Statistical Genetics Workshop in 2023, which was helpful for his training in psychiatric genetics. The Iowa Institute of Human Genomics performed hybridization of the beadchips of the replication cohort. JAW was also supported by the National Institute on Drug Abuse (R01DA052953), Department of Veterans Affairs (Merit Award, IO1BX004440), and Roy J. Carver Charitable Trust. We also thank Mr. Christopher Busby for text editing assistance. The content is solely the responsibility of the authors and does not necessarily represent the official views of the National Institutes of Health, the American Foundation for Suicide Prevention, the Department of Veterans Affairs, or the Roy J. Carver Charitable Trust.

## Notes

### Competing Interest Statement

The authors have declared no competing interest.

### Author Declarations

Ethics committee/IRB of the University of Texas Health Science Center gave ethical approval for this work (HSC-MS-09-0340). Ethics committee/IRB of the University of Iowa gave ethical approval for this work (201708703).

