## Supplementary Figures S1-S9 for "Blood epigenome-wide association studies of suicide attempt in adults with bipolar disorder"

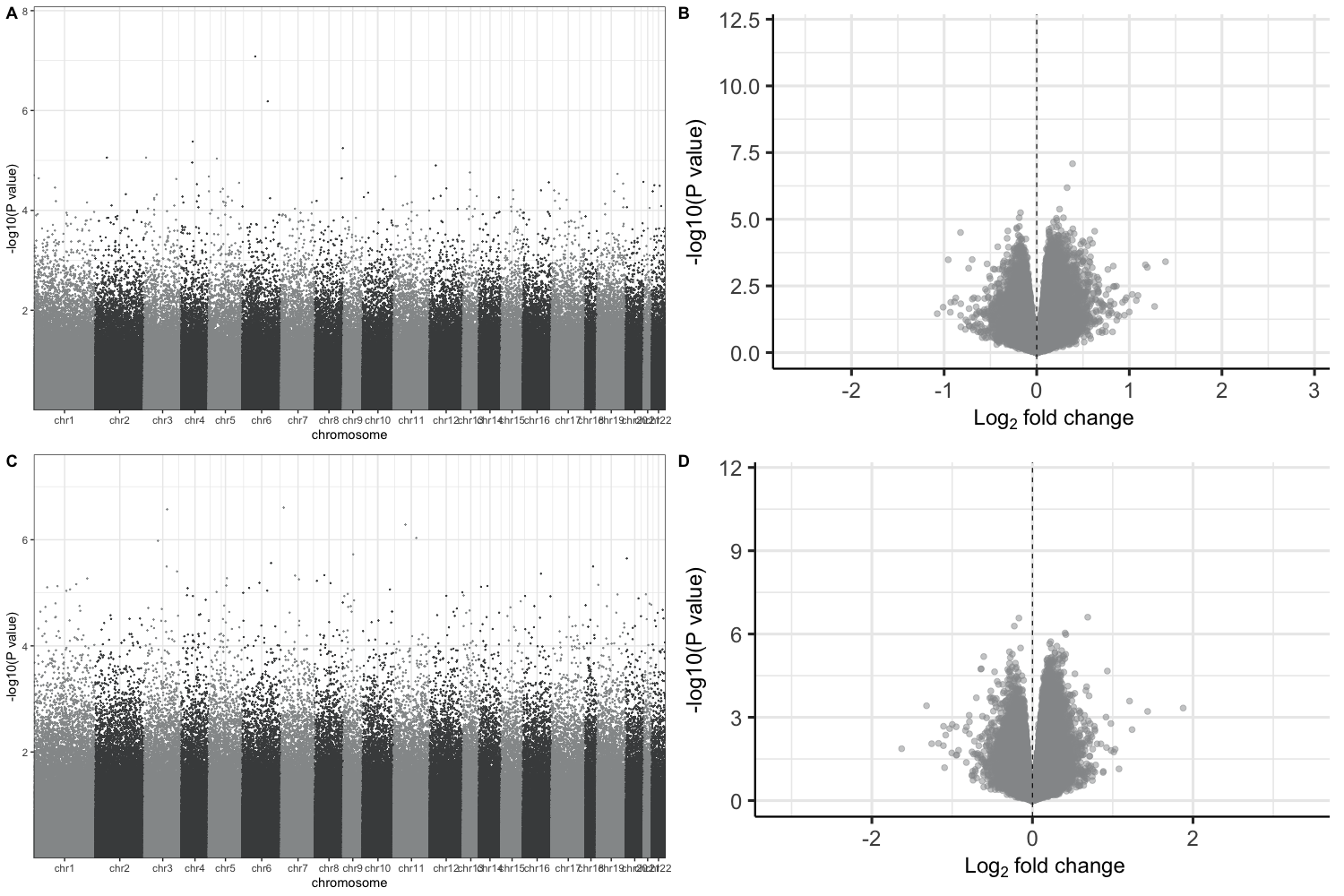

**Supplementary Figure S1. A** shows the Manhattan plot for the BD/SA vs. CON EWAS, **B** shows the Volcano plot for the BD/SA vs. CON EWAS, **C** shows the Manhattan plot for the BD/non-SA vs. CON EWAS, and **D** shows the Volcano plot for the BD/non-SA vs. CON EWAS. None of the differentially methylated positions passed false discovery rate (FDR) *q* < 0.05. BD - bipolar disorder; CON - controls; EWAS - epigenome-wide association study; FDR - false discovery rate; SA - suicide attempt.

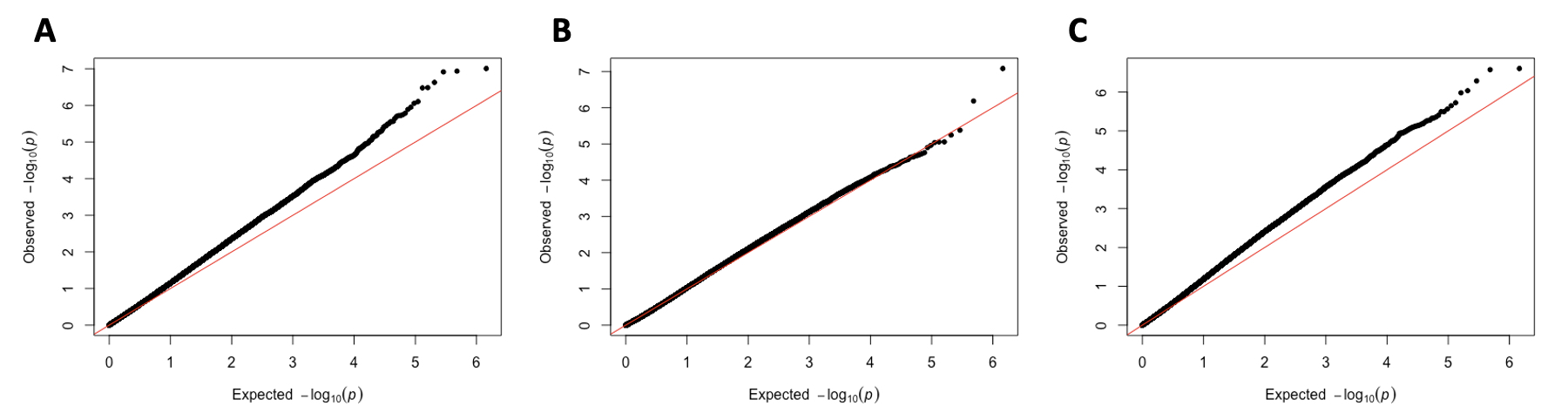

**Supplementary Figure S2.** Quantile-quantile (QQ) plots for the discovery cohort EWAS. **A** shows the QQ plot for the BD/SA vs. BD/non-SA EWAS (lambda inflation factor = 1.20), **B** shows the QQ plot for the BD/SA vs. CON EWAS (lambda inflation factor = 0.90), and **C** shows the QQ plot for the BD/non-SA vs. CON EWAS (lambda inflation factor = 1.21). BD - bipolar disorder; CON - controls; EWAS - epigenome-wide association study; SA - suicide attempt.

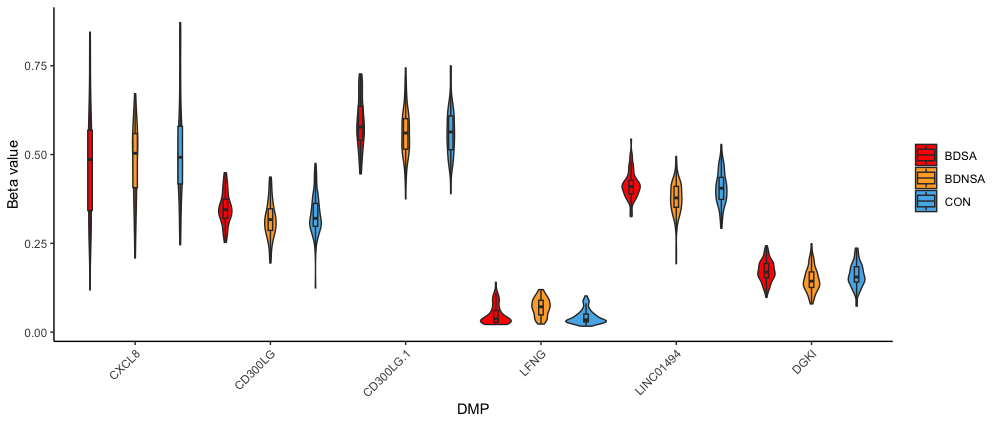

**Supplementary Figure S3.** Beta-value methylation levels for each FDR-significant DMP from the BD/SA vs. BD/non-SA contrast (labeled by annotated gene, most significant on left) between BD/SA, BD/non-SA, and CON groups demonstrate that there is not a consistent stepwise pattern in methylation associated with stepwise changes in severity (CON -> BD/non-SA -> BD/SA). This could suggest nonlinear/discontinuous methylation differences between groups. *CXCL8* - cg20244265; *CD300LG* - cg25876840; *CD300LG.1* - cg11476866; *LFNG* - cg15653194; *LINC01494* - cg00306112; *DGKI* - cg20242392. BD - bipolar disorder; CON - controls; DMP - differentially methylated positions; FDR - false discovery rate; SA - suicide attempt.

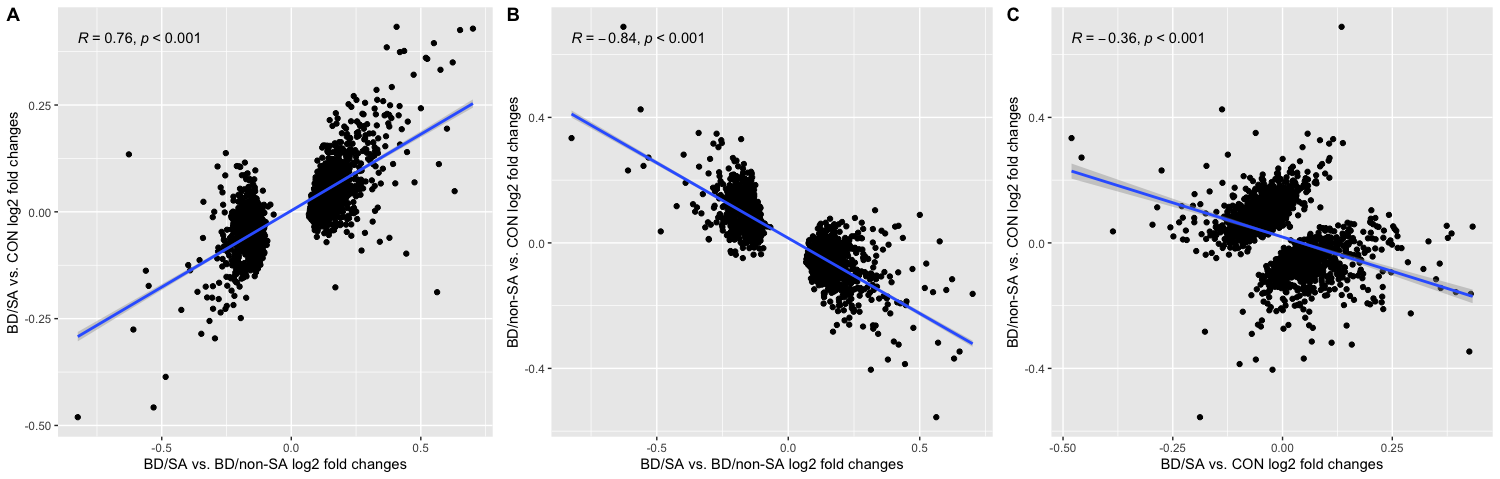

**Supplementary Figure S4.** Correlations among log2 fold changes for the three EWAS in the discovery cohort, focused on the DMPs at *p* < 0.001 in the BD/SA vs. BD/non-SA EWAS. **A** shows a strong positive correlation among log2 fold changes between BD/SA vs. BD/non-SA and BD/SA vs. CON EWAS, suggesting similar epigenetic patterns distinguishing BD/SA from BD/non-SA and BD/SA from CON. **B** shows a strong negative correlation between BD/SA vs. BD/non-SA and BD/non-SA vs. CON EWAS, suggesting differing patterns distinguishing BD/SA from BD/non-SA and BD/non-SA from CON and possibly indicating that BD/SA is not an exacerbation of BD/non-SA pathophysiology relative to CON. **C** shows a weak negative correlation between BD/SA vs. CON and BD/non-SA vs. CON EWAS, again suggesting that the differentiation between BD/SA and CON, and BD/non-SA and CON, is separable at the DMPs, which is expected given the results of the main EWAS. BD - bipolar disorder; CON - controls; DMP - differentially methylated probes, EWAS - epigenome-wide association study.

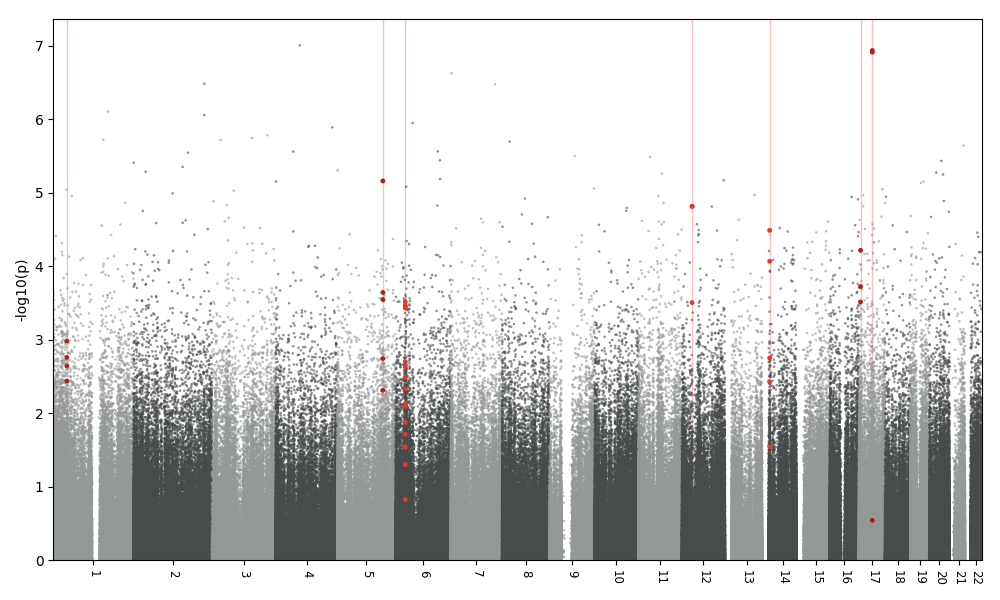

**Supplementary Figure S5.** Manhattan plot for DMRs, with Šidák-significant DMRs highlighted by red vertical lines, where individual points highlighted in red represent the associated CpG probes. The -log10 *p*-value for each individual probe can be seen on the y-axis. DMR - differentially methylated region.

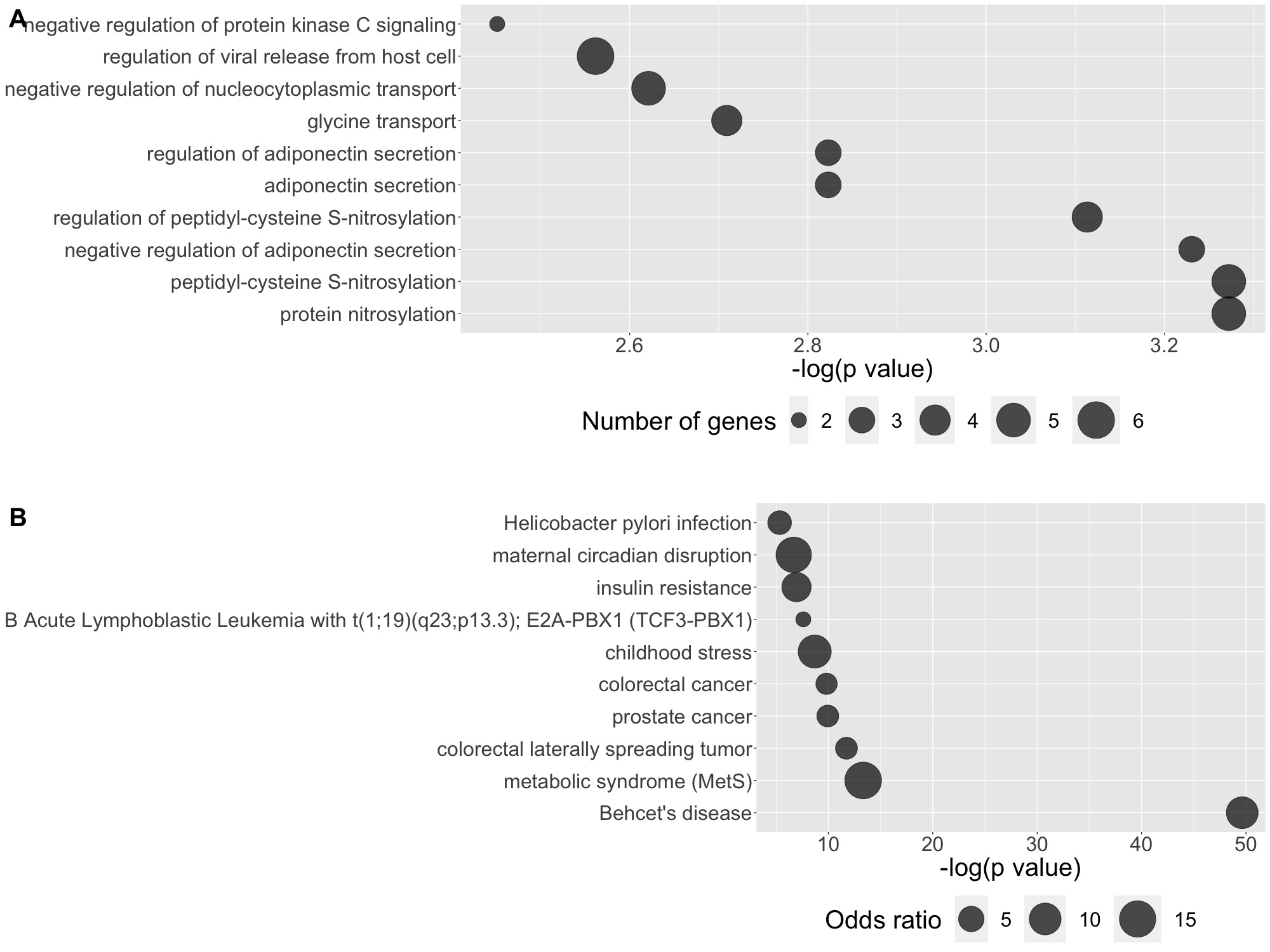

**Supplementary Figure S6.** Gene ontology and trait enrichment results for the discovery cohort BD/SA vs. CON EWAS using the DMPs at nominal *p* < 0.001. **A)** shows the top ten gene ontology pathways ordered by -log(p value), with the number of genes assigned to each pathway represented by the size of the point. **B)** shows the top ten traits ordered by -log(p value), with the odds ratio for each trait represented by the size of the point.

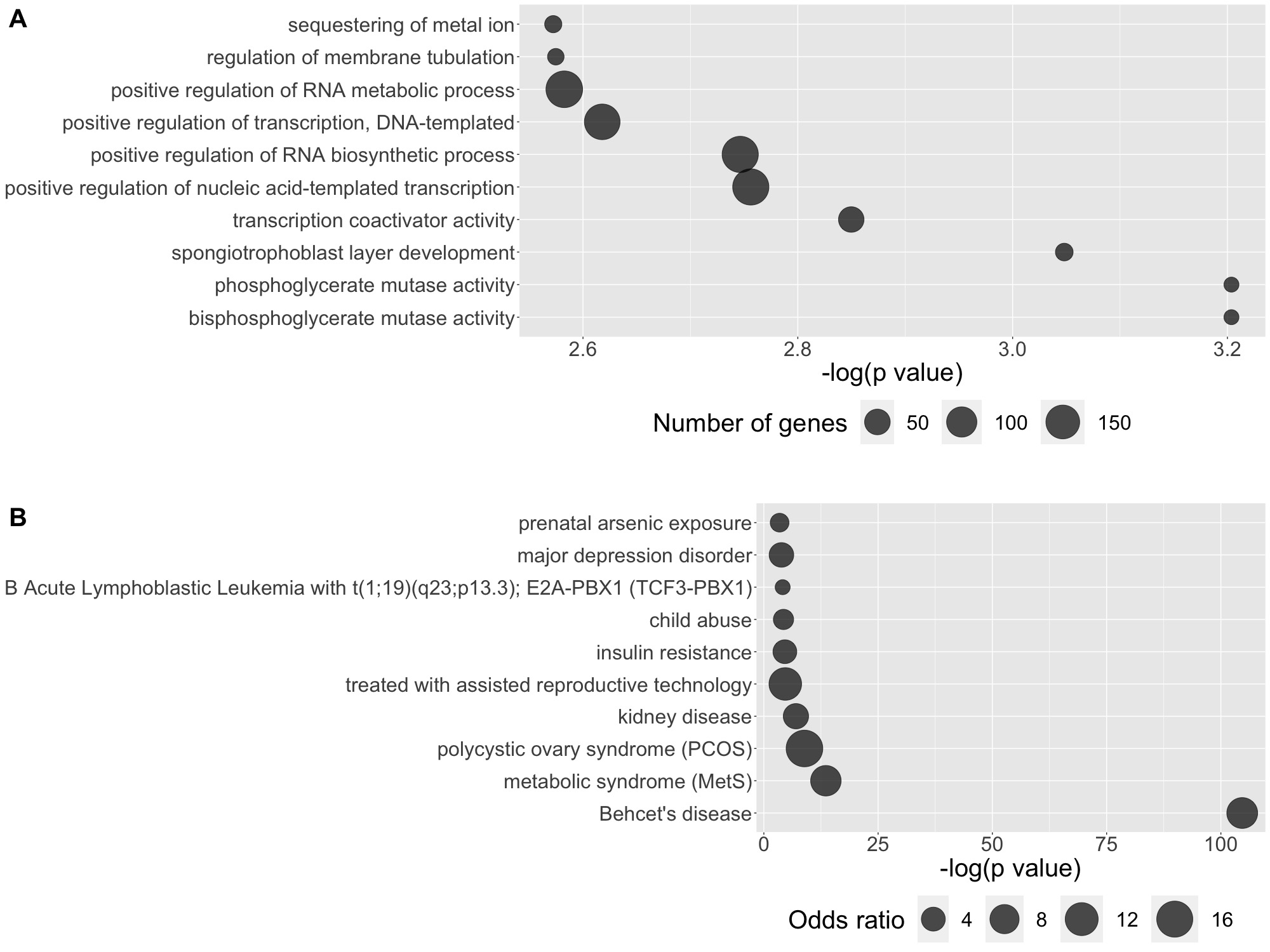

**Supplementary Figure S7.** Gene ontology and trait enrichment results for the discovery cohort BD/non-SA vs. CON EWAS using the DMPs at nominal *p* < 0.001. **A)** shows the top ten gene ontology pathways ordered by -log(p value), with the number of genes assigned to each pathway represented by the size of the point. **B)** shows the top ten traits ordered by -log(p value), with the odds ratio for each trait represented by the size of the point.

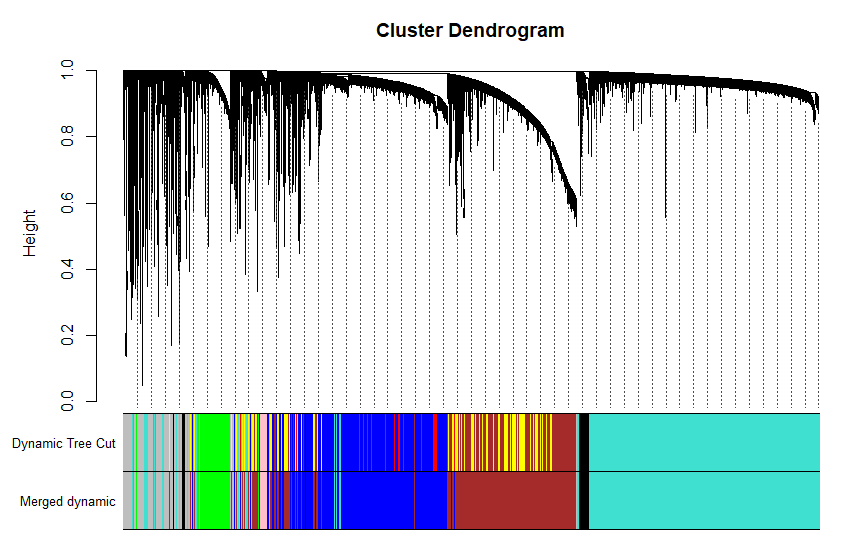

**Supplementary Figure S8.** Cluster dendrogram for the weighted gene co-methylation network analysis (WGCNA).

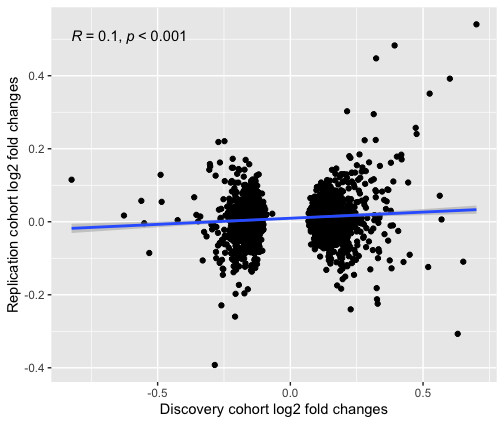

**Supplementary Figure S9.** Attempt to replicate patterns of log2 fold changes in the BD/SA vs. BD/non-SA contrast across the discovery and replication cohorts. Scatter plot of the log2 fold changes for the 1,958 discovery cohort sites with a nominal *p* < 0.001 (missing 114 probes which failed quality control in replication cohort) across both cohorts, with the blue line showing the linear fit (*r* = 0.10). BD - bipolar disorder; SA - suicide attempt.
